# Default mode network tau predicts future clinical decline in atypical early Alzheimer’s disease

**DOI:** 10.1101/2024.04.17.24305620

**Authors:** Yuta Katsumi, Inola A. Howe, Ryan Eckbo, Bonnie Wong, Megan Quimby, Daisy Hochberg, Scott M. McGinnis, Deepti Putcha, David A. Wolk, Alexandra Touroutoglou, Bradford C. Dickerson

## Abstract

Identifying individuals with early stage Alzheimer’s disease (AD) at greater risk of steeper clinical decline would allow professionals and loved ones to make better-informed medical, support, and life planning decisions. Despite accumulating evidence on the clinical prognostic value of tau PET in typical late-onset amnestic AD, its utility in predicting clinical decline in individuals with atypical forms of AD remains unclear. In this study, we examined the relationship between baseline tau PET signal and the rate of subsequent clinical decline in a sample of 48 A^+^/T^+^/N^+^ patients with mild cognitive impairment or mild dementia due to AD with atypical clinical phenotypes (Posterior Cortical Atrophy, logopenic variant Primary Progressive Aphasia, and amnestic syndrome with multi-domain impairment and age of onset < 65 years). All patients underwent structural magnetic resonance imaging (MRI), tau (^18^F-Flortaucipir) PET, and amyloid (either ^18^F-Florbetaben or ^11^C-Pittsburgh Compound B) PET scans at baseline. Each patient’s longitudinal clinical decline was assessed by calculating the annualized change in the Clinical Dementia Rating Sum-of-Boxes (CDR-SB) scores from baseline to follow-up (mean time interval = 14.55 ± 3.97 months). Our sample of early atypical AD patients showed an increase in CDR-SB by 1.18 ± 1.25 points per year: *t*(47) = 6.56, *p* < .001, *d* = 0.95. These AD patients showed prominent baseline tau burden in posterior cortical regions including the major nodes of the default mode network, including the angular gyrus, posterior cingulate cortex/precuneus, and lateral temporal cortex. Greater baseline tau in the broader default mode network predicted faster clinical decline. Tau in the default mode network was the strongest predictor of clinical decline, outperforming baseline clinical impairment, tau in other functional networks, and the magnitude of cortical atrophy and amyloid burden in the default mode network. Overall, these findings point to the contribution of baseline tau burden within the default mode network of the cerebral cortex to predicting the magnitude of clinical decline in a sample of atypical early AD patients one year later. This simple measure based on a tau PET scan could aid the development of a personalized prognostic, monitoring, and treatment plan tailored to each individual patient, which would help clinicians not only predict the natural evolution of the disease but also estimate the effect of disease-modifying therapies on slowing subsequent clinical decline given the patient’s tau burden while still early in the disease course.

## Introduction

Alzheimer’s disease (AD) is defined neuropathologically by the presence of β-amyloid plaques (Aβ) and neurofibrillary tangles composed of hyperphosphorylated tau^1^. The aggregation of these pathological proteins is thought to play a pivotal role in a neurodegenerative cascade leading to a phenotypically heterogeneous cognitive-behavioral dementia syndrome. With the advent of positron emission tomography (PET) radiotracers specifically binding to aggregated Aβ plaques or hyperphosphorylated tau, it is now possible to visualize and quantify these neuropathological features in living human brains, thus providing *in vivo* diagnostic confirmation. Tau PET in particular has garnered attention in recent years given converging evidence showing that the topography of elevated tau PET signal is more closely associated with co-localized neurodegeneration and the types and severity of symptoms than amyloid PET across the AD syndromic spectrum^2–10^.

While the utility of tau PET for diagnosis is established (with FDA approval in 2020), evidence has recently begun accruing to demonstrate its clinical prognostic value in early symptomatic AD. In patients with typical MCI or mild dementia due to AD, those with relatively greater cerebral tau PET signal show a relatively faster decline in cognitive test performance on the Mini Mental State Examination^11–17^, Addenbrooke’s Cognitive Examination^18^, or neuropsychological composite scores^11,14,15,19,20^. Given the importance of the Clinical Dementia Rating (CDR) scale as a clinical outcome measure in therapeutic trials and other studies (capturing both cognitive and functional impairment), surprisingly little work has been done on tau-based prognostication of longitudinal CDR change. One study of a mixed sample including asymptomatic, early stage AD, and moderate stage AD dementia patients showed that relatively greater tau PET signal predicted faster decline on the CDR Sum-of-Boxes (CDR-SB)^21^, while another study of early AD did not find a relationship between baseline tau PET and longitudinal CDR-SB change^22^. Of these studies, those that directly compared the prognostic utility of tau PET to that of other imaging (e.g., atrophy, amyloid PET) and/or fluid biomarkers of AD at baseline have consistently found that tau PET was the strongest predictor of decline on cognitive test measures^11–14,16–20^. A comparison of the prognostic utility of tau PET vs. amyloid PET and atrophy has not yet been performed with CDR-SB as an outcome measure. Furthermore, most of these studies have focused on examining individuals presenting with typical late-onset amnestic AD, leaving unclear the utility of tau PET as a prognostic tool in early symptomatic individuals with atypical features either based on non-amnestic clinical phenotype or young age of onset. We and others have shown that the topography of abnormal tau accumulation in atypical AD clinical phenotypes is spatially distinct from what is commonly observed with typical amnestic AD^7,9,10,23–26^. This suggests that definitions of regional tau deposition commonly used in studies primarily investigating amnestic late-onset AD (e.g., based on the Braak-staging scheme of tau progression^27,28^) would not be suitable for atypical AD.

One potential solution to this problem concerns the characterization of regional tau accumulation on the basis of large-scale functional networks of the brain^29,30^. Converging multimodal neuroimaging evidence in humans provides support for the hypothesis that pathological tau proteins spread through neuronal network connectivity in the cerebral cortex^24,31–36^. We have recently shown that individuals with atypical AD—across heterogeneous clinical phenotypes— consistently exhibit abnormal tau accumulation in the posterior nodes of the default mode network, including the posterior cingulate cortex (PCC), precuneus, and lateral parietal and temporal cortices bilaterally^25^. This indicates that, while the primary drivers of the earliest symptoms in atypical AD phenotypes are observed in other domain-specific networks (e.g., language, visuospatial)^24,37^, the default mode network may be commonly affected across the AD clinical spectrum. As early symptomatic AD progresses, tau spreads to prefrontal cortex, including nodes of the default mode network^26,38^. Thus, one plausible hypothesis is that individuals with atypical AD who exhibit tau accumulation that is more widespread and/or larger in magnitude within the default mode network would be likely to experience faster clinical decline.

In the present study, we sought to investigate the utility of in vivo tau pathology measures (^18^F-Flortaucipir [FTP] PET) localized to specific cortical functional networks in predicting subsequent clinical decline using the CDR-SB in a sample of patients with MCI and mild dementia due to AD (“early AD”) with a variety of clinical phenotypes of atypical AD. We additionally assessed in the same sample of patients the utility of regional cortical atrophy and amyloid PET for the same purpose, and compared the contributions of these imaging biomarkers to predicting cognitive and functional decline relative to that of FTP PET. On the basis of evidence reviewed above, we hypothesized that greater tau accumulation within the default mode network at baseline would predict faster longitudinal cognitive and functional decline. We also hypothesized that tau burden within the default mode network predicts subsequent clinical decline more strongly than cortical thickness or amyloid burden.

## Materials and methods

### Participants

The present study included 48 individuals with a clinical diagnosis of MCI or mild dementia due to AD with positive amyloid, tau, and neurodegeneration (A^+^/T^+^/N^+^) imaging biomarkers^39^, all of whom were recruited from the Massachusetts General Hospital (MGH) Frontotemporal Disorders Unit; see **Table 1** for demographic and clinical characteristics of the sample. This sample of early AD patients (mean Clinical Dementia Rating scale sum of boxes [CDR-SB] at baseline = 3.5 ± 1.87) mostly included atypical clinical phenotypes, consisting of individuals fulfilling diagnostic criteria for Posterior Cortical Atrophy^40–42^ (PCA) (*n* = 16), logopenic variant Primary Progressive Aphasia^43^ (lvPPA) (*n* = 15), amnestic syndrome with multi-domain impairment including dysexecutive AD (*n* = 17) with young age of onset (< 65 years)^44^. In addition, we included a sample of amyloid-negative (Aβ-) cognitively unimpaired control participants, whose data were used as a reference for quantifying elevated FTP uptake and cortical atrophy in our patients.

**Table 1.** Demographic and clinical characteristics of the sample.

| | AD | Aβ- CU |
| --- | --- | --- |
| *n* | 48 | 24 |
| Age (years) | 64.1 ± 8.16 | 67.62 ± 4.68 |
| Sex (M/F) | 20/28 | 12/12 |
| Education (years) | 16.7 ± 2.46 | <sup>a</sup>15.7 ± 2.25 |
| MMSE at baseline | <sup>b</sup>22.94 ± 4.99 | - |
| CDR at baseline | CDR 0 (*n* = 3)<br>CDR 0.5 (*n* = 28)<br>CDR 1 (*n* = 17) | CDR 0 (*n* = 24) |
| CDR-SB at baseline | 3.5 ± 1.87 | 0 |
| Baseline to follow-up (months) | 14.55 ± 3.97 | - |
| Neocortical composite amyloid (CL) | 96.32 ± 25.29 | 2.12 ± 6.48 |

<sup>a</sup>Data based on *n* = 13.
<sup>b</sup>Data based on *n* = 47; MMSE data were not available for a subset of patients (*n* = 21) and thus were estimated from their raw MoCA scores using the published conversion table<sup>52</sup>.
MMSE = Mini-Mental State Examination; CDR = Clinical Dementia Rating; CDR-SB = Clinical Dementia Rating Sum-of-Boxes; CL = Centiloid

All patients received a standard clinical evaluation comprising a structured history obtained from both patient and informant to inform clinician scoring on the CDR, comprehensive neurological and psychiatric history and exam, and neuropsychological assessment. Clinical diagnostic formulation was performed through consensus conference by our multidisciplinary team of neurologists, neuropsychologists, and speech and language pathologists, with each patient classified based on all available clinical information as having a 3-step diagnostic formulation of mild cognitive impairment or dementia (Cognitive Functional Status), a specific Cognitive-Behavioral Syndrome, and a likely etiologic neuropathologic diagnosis^45^. Each patient’s longitudinal clinical decline was measured by calculating the annualized change in the CDR-SB scores from baseline to approximately one year later. At baseline, all patients underwent neuroimaging sessions involving structural MRI, FTP PET, and amyloid (either ^18^F-Florbetaben [FBB] or ^11^C-Pittsburgh Compound B [PiB]) PET scans. Aβ positivity was determined by a combination of visual read and mean amyloid PET signal extracted from a cortical composite region of interest according to previously published procedures^46–48^. Determination of tau and neurodegeneration positivity was conducted by visual read using internal methods similar to published work^49–51^.

We also included a group of Aβ-cognitively unimpaired individuals, all of whom performed within normal limits on neuropsychological testing, had normal brain structure based on MRI, and low cerebral amyloid based on quantitative analysis of PiB PET data (FLR DVR < 1.2). This control sample was used as a reference for quantifying elevated signal in FTP PET and cortical thickness in early AD patients. Individuals were excluded from our patient and control groups if they had a primary psychiatric or other neurologic disorder including major cerebrovascular infarct or stroke, seizure, brain tumor, hydrocephalus, multiple sclerosis, HIV-associated cognitive impairment, or acute encephalopathy. This work was carried out in accordance with The Code of Ethics of the World Medical Association (Declaration of Helsinki) for experiments involving humans. All participants and their informants/caregivers provided informed consent in accordance with the protocol approved by the MassGeneral Brigham HealthCare System Human Research Committee Institutional Review Board in Boston, Massachusetts.

### Neuroimaging data acquisition and preprocessing

All patients underwent structural MRI, FTP PET, and amyloid PET scans at baseline. Structural MRI data were acquired from each participant on either a 3 Tesla Siemens Prisma Fit scanner using a T1-weighted magnetization prepared rapid acquisition sequence (MPRAGE) (repetition time [TR] = 2300 ms, echo time [TE] = 2.98 ms, flip angle = 9°, slice thickness = 1 mm, field of view [FOV] = 240 × 256 mm^2^) or on a 3 Tesla Siemens Tim Trio using a multi-echo MPRAGE sequence (TR = 2530 ms, TEs = 1.64/3.5/5.36/7.22 ms, flip angle = 7°, slice thickness = 1 mm, FOV = 256 × 256 mm^2^). Each participant’s (ME)MPRAGE data underwent intensity normalization, skull stripping, and an automated segmentation of cerebral white matter to locate the gray matter/white matter boundary via FreeSurfer v6.0 (https://surfer.nmr.mgh.harvard.edu). Defects in the surface topology were corrected^53^, and the gray/white boundary was deformed outward using an algorithm designed to obtain an explicit representation of the pial surface. We visually inspected each participant’s cortical surface reconstruction for technical accuracy and manually edited it when necessary. Cortical thickness was calculated as the closest distance from the gray/white boundary to the gray/CSF boundary at each vertex on the tessellated surface^54^. For the purpose of Centiloid scaling only, we additionally processed each patient’s (ME)MPRAGE data via FreeSurfer v7.1 to keep a consistency with the current ADNI approach^55^.

PET data were acquired using a Siemens (Knoxville, TN) ECAT HR + scanner. Tau PET images were acquired from 80 to 100 min after the bolus injection of ∼10.0 mCi of FTP (4 × 5 min frames). Amyloid PET data were acquired either from 90 to 110 min after the injection of ∼8 mCi of FBB (4 × 5 min frames) or a 60 min dynamic acquisition after the injection of 8.5 to 10.5 mCi of PiB (69 frames; 12 × 15 sec followed by 57 × 60 sec). All PET data were reconstructed and attenuation corrected; each frame was evaluated to verify adequate count statistics and interframe head motion was corrected. Individual FTP PET frames were aligned to the first via FSL FLIRT^56^ and were averaged. Further processing of FTP PET data were performed via the PetSurfer tools^57,58^. Each participant’s FTP PET data were first rigidly co-registered to their anatomical volume and the accuracy of cross-modal spatial registration was confirmed by visual inspection. FTP PET data were then corrected for partial volume effects. Specifically, based on each participant’s high-resolution tissue segmentation derived by the standard Desikan-Killiany atlas^59^, the symmetric geometric transfer matrix method was used to correct for spill-in and spill-out effects between adjacent brain tissue types, with a point spread function of 6 mm^57,58^. Using partial volume-corrected data, we derived the FTP standard uptake value ratio (SUVR) image per participant with whole cerebellar gray matter as a reference region. Finally, FTP SUVR maps were resampled to *fsaverage* space and smoothed geodesically with FWHM of 8 mm.

### Calibration of amyloid PET SUVR data to Centiloid units

In the current patient sample, baseline amyloid deposition was quantified using FBB PET for 21 patients, whereas PiB PET was used for 27 patients. To harmonize the data across these different radioligands, we converted both FBB and PiB SUVR images to Centiloid (CL) units following the published guidelines^55,60^. See **Supplementary Methods** and **Supplementary** Fig. 1 for details.

### Statistical analysis

Using individual FTP SUVR maps as inputs, we first constructed a whole-cortex vertex-wise general linear model (GLM) using FreeSurfer’s *mri_glmfit* function to identify areas of the cerebral cortex where early AD patients showed greater FTP uptake than the Aβ-control participants. Statistical significance was assessed with a vertex-wise threshold of *p* < 10^-8^ uncorrected for multiple comparisons. To characterize the spatial topography of abnormal FTP signal in early AD with respect to large-scale functional networks of the cerebral cortex, we utilized an established parcellation with seven spatially non-overlapping networks (visual, somatomotor, dorsal attention, ventral attention, limbic, frontoparietal, and default)^30^. Here, we adhere to the original and conventional use of these networks, although we acknowledge that the “default” and “limbic” networks are not always distinguished in the literature^61^ and that both networks contain agranular, limbic tissue^62,63^. Given our a priori hypotheses about the default mode network, we additionally characterized FTP uptake separately for each cortical lobar region of interest (ROI) within this network (frontal, parietal, medial temporal, and lateral temporal).

Next, we constructed a vertex-wise GLM to examine the bivariate association between baseline FTP uptake and annualized change in CDR-SB across all AD patients. Statistical significance was assessed with a vertex-wise threshold of *p* < .01 uncorrected for multiple comparisons. We additionally compared the magnitude of correlation across the four ROIs within the default mode network by extracting their mean FTP uptake. We then performed a series of multiple linear regression analyses with annualized change in CDR-SB as the outcome variable, while using mean FTP uptake in each functional network as the main predictor in separate models (i.e., *simple* network models), while controlling for age, sex, and CDR-SB at baseline as covariates of no interest. Performance of regression models was evaluated using adjusted *R*^2^ as well as changes in Akaike information criterion (AIC) values, where ΔAIC < -2 was considered evidence for significant improvement in model fit^17,64^. Following the examination of simple network models, we investigated the complementary contributions of suprathreshold functional networks to predicting CDR-SB change via an automated data-driven model selection using the *MuMIn* package (v1.47.5) run in *R* (v4.2.1)^65^. This analysis evaluated the performance of models constructed using all possible variable combinations and ranking them based on their AIC, with the goal to identify the most parsimonious model defined as the one with the fewest predictors within two AIC points from the lowest AIC^66^.

Having established the specific role of tau in the default mode network in predicting future clinical decline, we next investigated the extent to which other major elements of AD neuropathologic features (i.e., cortical atrophy and amyloid burden) have complementary roles in prognostication. To this end, we first examined baseline maps of cortical thickness and amyloid PET using vertex-wise GLMs to identify areas of abnormal signal as well as the topography and magnitude of association with annualized CDR-SB change in early AD patients. We then constructed a series of multiple linear regression models to assess complementary contributions of tau, cortical thickness, and amyloid in the default mode network to predicting CDR-SB change. Finally, as a first step toward developing a tool for individualized risk stratification, we conducted an exploratory analysis to see whether a subset of patients exhibiting faster clinical decline with the rest of the patient sample (i.e., CDR-SB change >1 *SD* greater than the group mean) would differ on baseline tau burden, cortical thickness, or amyloid burden. Unless otherwise specified, statistical significance for group-level statistical analyses was assessed at *p* < .05.

## Results

### Early AD is characterized by cortical tau deposition in multiple cerebral functional networks

A vertex-wise comparison of FTP uptake between AD patients and Aβ-control participants revealed elevated signal predominantly in posterior cortical regions with modest lateral prefrontal involvement. These regions overlapped substantially with the major nodes of the default mode network, including the angular gyrus, posterior cingulate cortex/precuneus, and lateral temporal cortex, with lesser involvement of the medial temporal lobe (**Fig. 1A**). To further characterize the spatial topography of cortical tau deposition, we calculated the mean FTP SUVR across all vertices falling within the boundaries of each of the seven canonical functional networks^30^ as well as four lobar regions within the default mode network. Within the default mode network, the largest effect size of between-group differences was observed in the lateral temporal areas, followed by the parietal areas (**Fig. 1B**). Among the rest of the canonical functional networks, the dorsal attention network showed the largest effect size of group differences in mean FTP uptake, with all the other networks showing smaller effect sizes than the default mode network (**Fig. 1C**).

**Figure 1.**
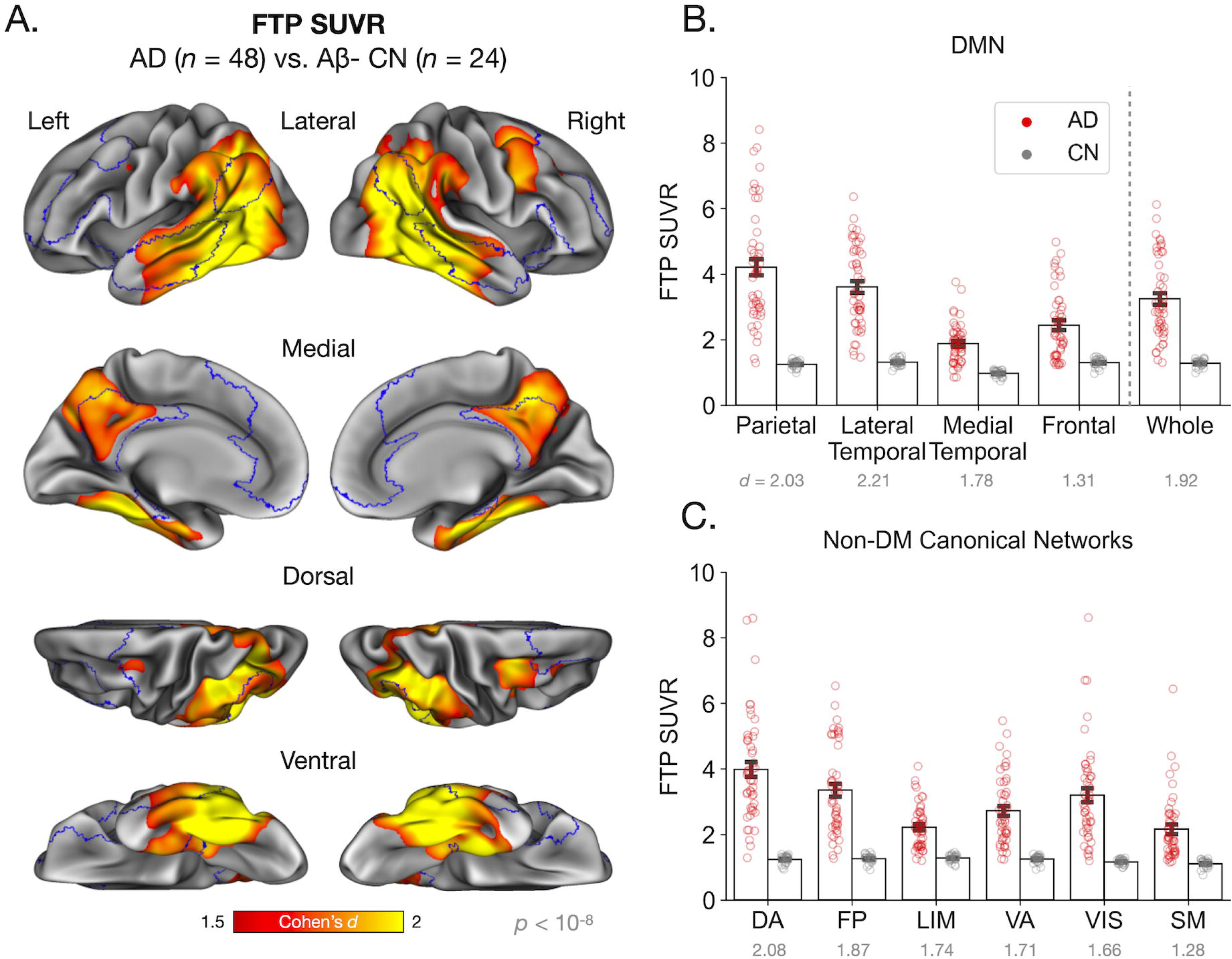
Mean ^18^F-Flortaucipir PET signal is high across the posterior default mode network in early A^+^/T^+^/N^+^ atypical Alzheimer’s disease. **(A)** Colored vertices on the cortical surface map indicate areas where A^+^T^+^N^+^ Alzheimer’s disease (AD) patients (*n* = 48) had greater ^18^F-Flortaucipir (FTP) SUVR than Aβ-cognitively unimpaired participants (*n* = 24). Statistical significance was assessed at vertex-wise *p* < 10^-8^. The default mode network (DMN) is outlined in blue. (B) Bar plot represents the mean FTP SUVR values calculated for each cortical lobar region of interest (ROI) part of the default mode network as well as the whole network separately for each group. These ROIs were defined based on a parcellation of the default mode network from Yeo et al.^30^. Error bars represent one standard error of the mean. Colored circles overlaid on the bar plot represent individual participants in each group. Values below *x* axis labels indicate Cohen’s *d* effect sizes. All group differences are statistically significant at *p* < .001. (C) Similar to (B) above, the bar plot here represents mean FTP SUVR values of the rest of the canonical functional networks of the cerebral cortex as defined by Yeo et al.^30^. All group differences are statistically significant at *p* < .001. DA = dorsal attention; FP = frontoparietal; VA = ventral attention; LIM = limbic; VIS = visual; SM = somatomotor.

### Greater tau burden in the default mode network at baseline predicts faster clinical decline in early AD

We used the CDR-SB to measure approximately 1-year clinical progression in these early AD patients. Each patient’s longitudinal clinical decline was assessed by calculating the annualized change in the CDR-SB scores from baseline to a follow-up visit (mean time interval = 14.55 ± 3.97 months). To control for individual variability in clinical follow-up, we computed the annualized change in CDR-SB scores in each patient ([CDR-SB at follow-up – CDR-SB at baseline]/time interval in years). Our early AD patients showed an increase in CDR-SB by 1.18 ± 1.25 points per year (*t*[47] = 6.56, *p* < .001, *d* = 0.95). We then performed a vertex-wise bivariate correlation analysis to examine the relationship between baseline FTP uptake and annualized change in CDR-SB across patients. This analysis revealed that baseline tau deposition in widespread areas of the cerebral cortex—which substantially overlapped with the default mode network—is associated with the magnitude of clinical decline over time (**Fig. 2A**). Unlike the analysis of group differences in baseline FTP uptake, the association between baseline FTP uptake and annualized CDR-SB increase was comparably strong across different lobar ROIs within this network (**Fig. 2B**). Based on this observation, our subsequent regression analyses focused on FTP uptake and other imaging biomarker data extracted from the whole network level.

**Figure 2.**
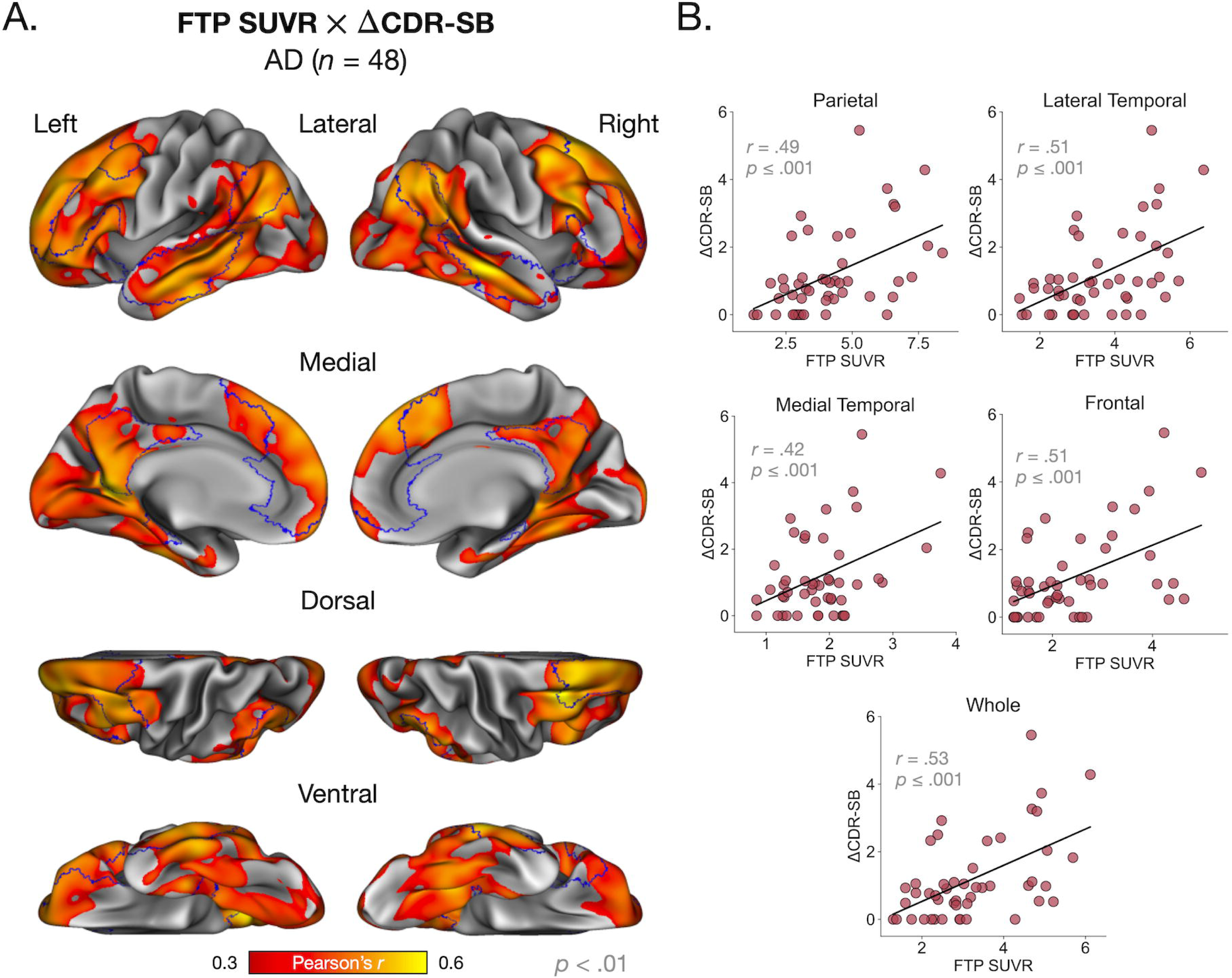
Greater baseline cerebral ^18^F-Flortaucipir PET signal in nearly the entire default mode network predicts faster subsequent clinical decline in early atypical AD. (**A**) Colored vertices on the cortical surface map indicate areas where greater FTP SUVR at baseline predicted faster clinical decline (as measured by the annualized change in CDR-SB scores) in patients with early AD (*N* = 48). Statistical significance was assessed at vertex-wise *p* < .01. Although the average baseline group elevation of tau is localized in the posterior default mode network (DMN) (Fig. 1A), this map shows that those individuals with more widespread tau in the DMN—including in not only posterior DMN but also anterior regions—progress faster. (**B**) Scatterplots depict zero-order bivariate associations between mean FTP SUVR extracted from lobar ROIs of the DMN as well as the entire network and the annualized change in CDR-SB scores. Consistent with vertex-wise results shown in (**A**), the pattern of associations is overall similar across ROIs, and shows that the overall tau burden throughout the DMN at baseline is a strong predictor of the rate of decline in the coming year.

### Baseline tau burden in the default mode network is a stronger predictor of clinical decline than that in other functional networks

**Table 2** summarizes the results of simple network models where we used FTP uptake in each functional network as the main predictor of annualized change in CDR-SB, while account for the effect of age, sex, and baseline CDR-SB as covariates. The *basic* model with age, sex, and CDR-SB at baseline revealed that higher CDR-SB scores at baseline predicted greater change in CDR-SB (*β* = 0.32, *SE* = 0.14, *p* ≤ .027). Controlling for these baseline measures, the inclusion of FTP uptake in the default mode network as a predictor in the model significantly improved fit (*F*(1,43) = 13.30, *p* < .001, ΔAIC = -10.94, Δ*R*^2^ = .20), such that greater FTP uptake in this network predicted greater change in CDR-SB (*β* = 0.54, *SE* = 0.15, *p* < .001). In addition, we found that modeling FTP uptake in several other functional networks resulted in significantly improved fit, including the frontoparietal (*F*(1,43) = 7.77, *p* ≤ .0023, ΔAIC = -8.54, Δ*R*^2^ = .16), limbic (*F*(1,43) = 6.68, *p* ≤ .013, ΔAIC = -4.93, Δ*R*^2^ = .11), ventral attention (*F*(1,43) = 5.63, *p* ≤ .022, ΔAIC = -3.91, Δ*R*^2^ = .09), and visual (*F*(1,43) = 4.76, *p* ≤ .035, ΔAIC = -3.04, Δ*R*^2^ = .07) networks. However, the model with FTP uptake in the default mode network was the strongest predictor of CDR-SB change compared with all the other simple network models. In general, baseline CDR-SB was a weaker predictor of CDR-SB change and was in many cases statistically insignificant (*p* > .05) when modeled with network-specific FTP uptake.

**Table 2.** Basic and simple network models predicting annualized change in CDR-SB in patients with atypical early AD.

| Model | Age | Sex | Baseline CDR-SB | Default | Frontoparietal | Visual | Limbic | Dorsal Attention | Ventral Attention | Somatomotor | Adjusted $R^2$ | $p$ | AIC |
| --- | --- | --- | --- | --- | --- | --- | --- | --- | --- | --- | --- | --- | --- |
| Basic | -0.07 (.63) | 0.46 (.11) | 0.32 (.027)* | - | - | - | - | - | - | - | .10 | .051 | -1.40 |
| Full | 0.09 (.60) | 0.14 (.61) | 0.18 (.21) | 0.64 (.32) | 0.09 (.90) | 0.95 (.004)* | -0.17 (.54) | -1.54 (.012)* | 0.97 (.13) | -0.24 (.39) | .36 | .0018* | -12.05 |
| **Default mode** | **0.18 (.21)** | **0.19 (.46)** | **0.24 (.062)** | **0.54 (<.001)\*** | - | - | - | - | - | - | **.30** | **<.001\*** | **-12.34** |
| Frontoparietal | 0.22 (.17) | 0.14 (.61) | 0.24 (.076) | - | 0.54 (.002)* | - | - | - | - | - | .26 | .0017* | -9.94 |
| Visual | -0.01 (.96) | 0.33 (.24) | 0.20 (.17) | - | - | 0.33 (.035)* | - | - | - | - | .17 | .015* | -4.44 |
| Limbic | -0.02 (.87) | 0.31 (.26) | 0.28 (.042)* | - | - | - | 0.35 (.013)* | - | - | - | .21 | .0072* | -6.33 |
| Dorsal Attention | 0.09 (.58) | 0.29 (.32) | 0.26 (.073) | - | - | - | - | 0.33 (.065) | - | - | .15 | .024* | -3.24 |
| Ventral Attention | 0.15 (.36) | 0.25 (.38) | 0.30 (.030)* | - | - | - | - | - | 0.40 (.022)* | - | .19 | .011* | -5.31 |
| Somatomotor | 0.02 (.90) | 0.43 (.13) | 0.29 (.046)* | - | - | - | - | - | - | 0.20 (.22) | .11 | .055 | -1.12 |

Predictor values indicate regression coefficients (p values). \* $P < .05$
The best performing model, defined as having the largest $\Delta AIC$ and $\Delta R^2$ compared with the basic model, is **bolded**, where FTP uptake in the default mode network is the only significant predictor. CDR-SB = Clinical Dementia Rating Sum-of-Boxes; AIC = Akaike information criterion.

### Data-driven model selection confirms the utility of tau in the default mode network in predicting clinical decline

Extending the simple network models reported above, we next sought to investigate complementary contributions of FTP uptake in different networks to predicting CDR-SB change. To this end, we conducted an automated data-driven model selection by testing all possible variable combinations and ranking the models based on their AIC^66^. In this analysis, we considered the five networks whose simple models provided better fit compared with the basic model, while controlling for age, sex, and baseline CDR-SB. Therefore, model selection was performed with a total of 256 models (255 based on different combinations of these predictors and the intercept-only model). This analysis revealed that the most parsimonious model—defined as the model with the fewest predictors within two AIC points from the lowest AIC—was the one with FTP uptake in the default mode network as the sole predictor (**Table 3**).

**Table 3.**
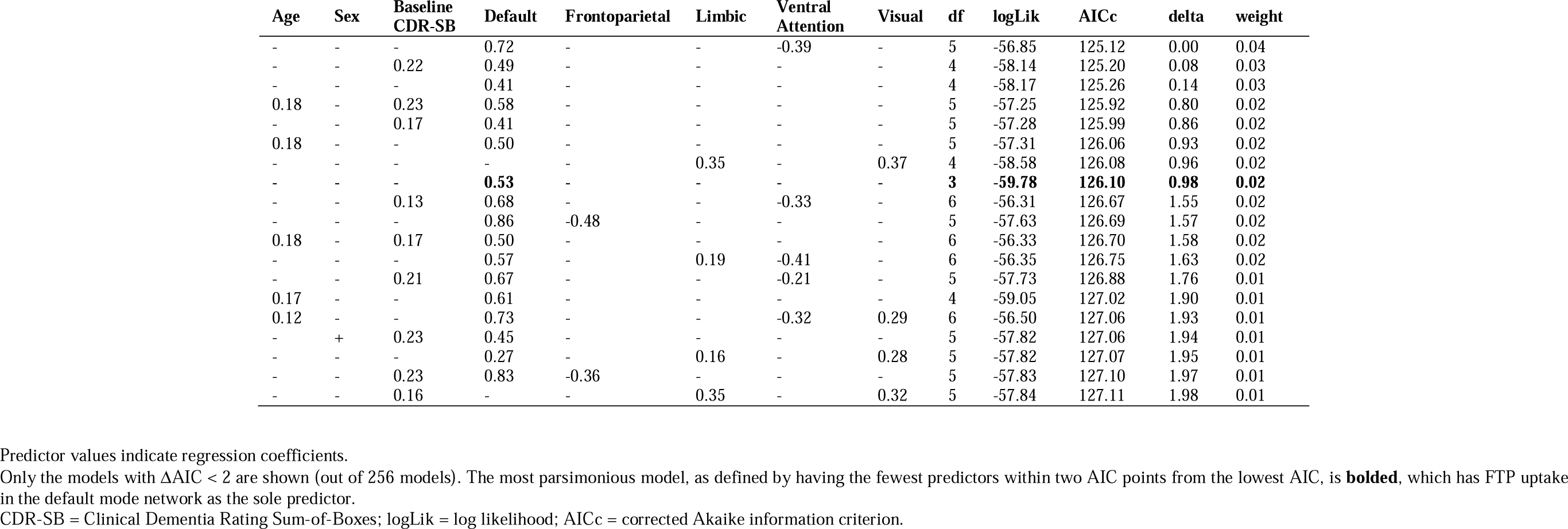
Selection of the most parsimonious model for predicting annualized change in CDR-SB in patients with atypical early AD.

### Complementary contributions of tau and neurodegeneration, but not amyloid, in the default mode network to predicting clinical decline

Following the analysis of FTP PET data, we examined baseline cortical thickness and amyloid PET data using vertex-wise GLMs to identify areas of abnormal signal as well as the topography and magnitude of association with annualized CDR-SB change in this early AD patient sample. We found prominent reduction in thickness (i.e., greater atrophy) that was co-localized with elevated FTP uptake in cortical areas including part of the default mode network, although of much lesser magnitude and spatial extent. Amyloid PET signal was prominently elevated in more widespread areas of the cerebral cortex, particularly in broader heteromodal association areas as well as limbic cortices (**Supplementary** Fig. 2). Again mirroring the pattern observed with FTP uptake, baseline cortical thickness in regions including those within the boundaries of the default mode network predicted subsequent clinical decline; however, this effect was both smaller in magnitude and narrower in spatial extent compared with what we observed with FTP uptake. The magnitude of the relationship between baseline amyloid PET signal and subsequent clinical decline was only sparsely observed (**Fig. 3**).

**Figure 3.**
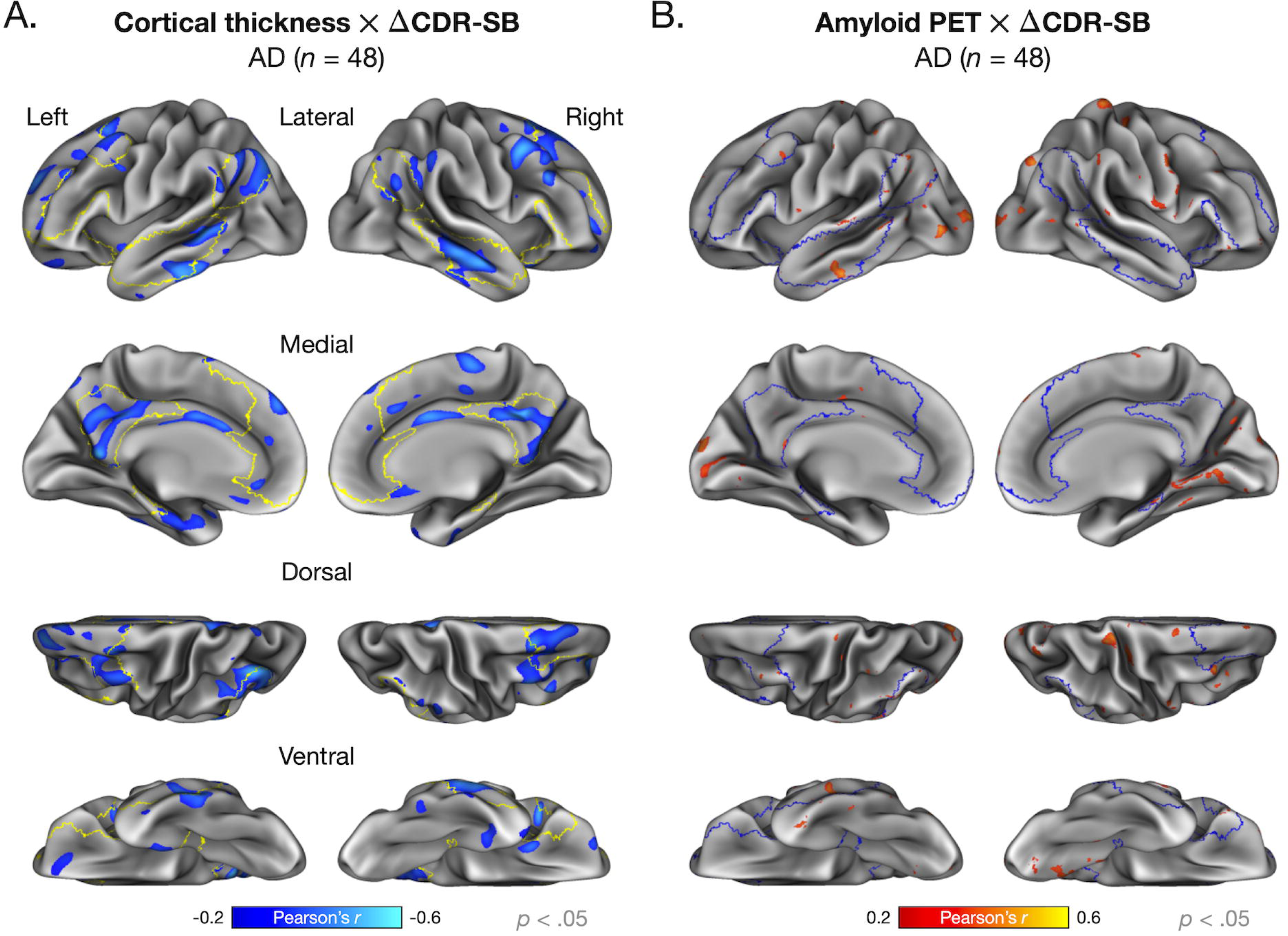
Association between baseline cortical thickness and amyloid deposition and subsequent clinical decline in atypical early AD. Colored vertices on the cortical surface maps indicate areas where (**A**) reduced cortical thickness or (**B**) greater amyloid deposition (expressed in Centiloid units) at baseline predicted faster clinical decline (as measured by the annualized change in CDR-SB scores) in patients with early AD (*N* = 48). Statistical significance was assessed at vertex-wise *p* < .05.

Extending vertex-wise analyses, we next constructed multiple linear regression models to predict CDR-SB change to investigate the contribution of individual biomarker data (FTP PET, cortical thickness, and amyloid PET) within the default mode network, controlling for age, sex, and baseline CDR-SB. We found that including cortical thickness of the default mode network as a predictor improved model fit (*F*(1,43) = 6.03, *p* ≤ .018, ΔAIC = -4.3, Δ*R*^2^ = .10), whereas amyloid PET did not (*F*(1,43) = 0.19, *p* ≤ .66, ΔAIC = 1.02, Δ*R*^2^ = -.01). The model with FTP uptake in the default mode network still yielded the largest effect size and lowest AIC value (**Table 4**). Finally, we conducted a data-driven model selection to identify the most parsimonious model given all possible variable combinations. This analysis showed that the model with two predictors—FTP uptake in the default mode network with either cortical thickness in the same network or baseline CDR-SB scores—as being the most parsimonious, with comparable model performance (**Table 5**).

**Table 4.**
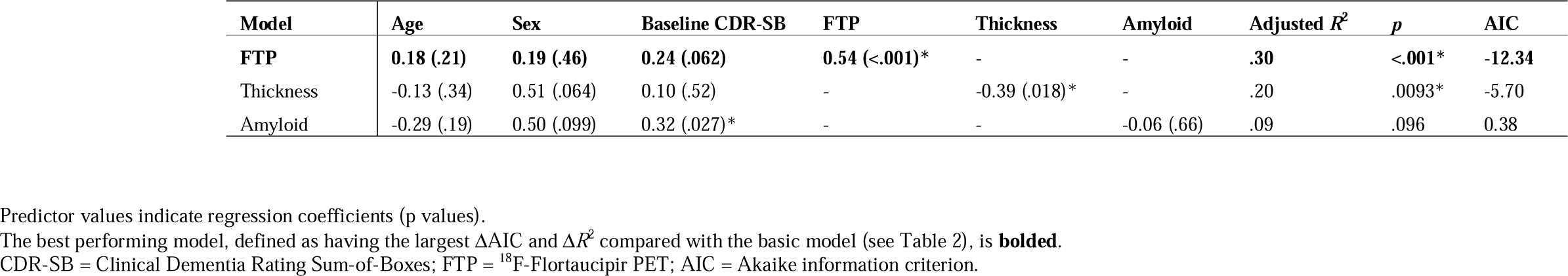
Multiple linear regression models predicting annualized change in CDR-SB in patients with atypical early AD by individual imaging biomarkers.

**Table 5.**
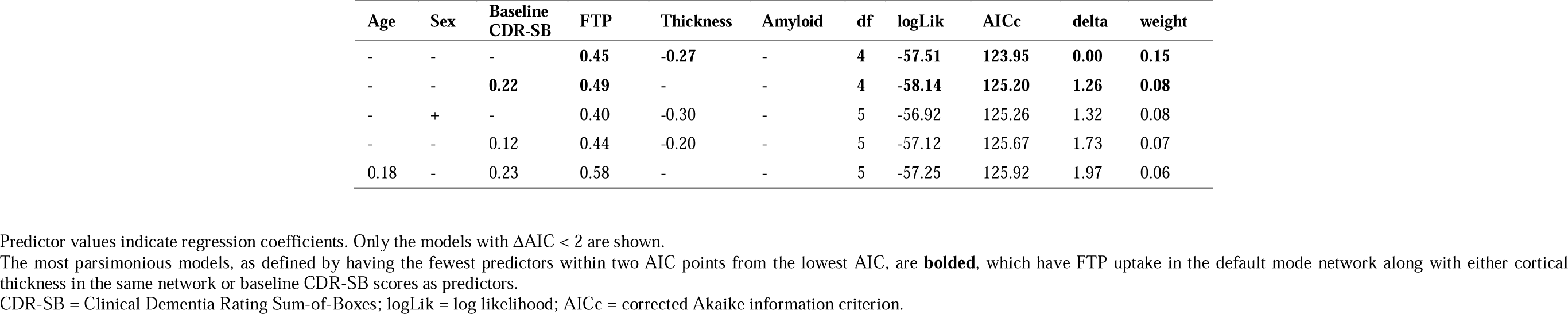
Selection of the most parsimonious multimodal model for predicting annualized change in CDR-SB in patients with atypical early AD.

| Age | Sex | Baseline CDR-SB | FTP | Thickness | Amyloid | df | logLik | AICc | delta | weight |
| --- | --- | --- | --- | --- | --- | --- | --- | --- | --- | --- |
| - | - | - | **0.45** | **-0.27** | - | **4** | **-57.51** | **123.95** | **0.00** | **0.15** |
| - | - | **0.22** | **0.49** | - | - | **4** | **-58.14** | **125.20** | **1.26** | **0.08** |
| - | + | - | 0.40 | -0.30 | - | 5 | -56.92 | 125.26 | 1.32 | 0.08 |
| - | - | 0.12 | 0.44 | -0.20 | - | 5 | -57.12 | 125.67 | 1.73 | 0.07 |
| 0.18 | - | 0.23 | 0.58 | - | - | 5 | -57.25 | 125.92 | 1.97 | 0.06 |

Predictor values indicate regression coefficients. Only the models with $\Delta AIC < 2$ are shown.
The most parsimonious models, as defined by having the fewest predictors within two AIC points from the lowest AIC, are **bolded**, which have FTP uptake in the default mode network along with either cortical thickness in the same network or baseline CDR-SB scores as predictors.
CDR-SB = Clinical Dementia Rating Sum-of-Boxes; logLik = log likelihood; AICc = corrected Akaike information criterion.

Finally, we identified a subset of early AD patients (*n* = 7) exhibiting faster clinical decline compared with the rest of the patient sample (i.e., CDR-SB change >1 *SD* greater than the group mean). Consistent with the regression results reported above, comparisons of these two patient subgroups revealed that faster clinical decline was associated with higher baseline CDR-SB scores (*t*(46) = 2.38, *p* ≤ .021) as well as greater FTP uptake (*t*(46) = 2.58, *p* ≤ .013) and reduced cortical thickness (*t*(46) = -2.55, *p* ≤ .014) in the default mode network, whereas the two groups did not differ in the composition of sexes (χ^2^ (1, *N* = 48) = 2.52, *p* ≤ .11), age (*t*(46) = -0.18, *p* ≤ .86), education (*t*(46) = -1.16, *p* ≤ .25), or amyloid deposition in this network (*t*(46) = 0.82, *p* ≤ .42) (**Supplementary Table 1** and **Supplementary** Fig. 2).

## Discussion

Patients at the early clinical stage of AD (MCI or mild dementia) are most commonly targeted in clinical trials focusing on the development of disease-modifying therapies, allowing for maximal opportunities to modulate the trajectory of cognitive and functional decline before substantial irreversible neurodegeneration and clinical impairment is present. Tau PET has already demonstrated important therapeutic relevance, as early symptomatic AD patients with relatively higher cortical tau accumulation are less likely to benefit from at least one anti-amyloid monoclonal antibody^67,68^. Clarifying the utility of tau burden in predicting future clinical decline would be an important first step toward implementing risk stratification strategies to identify patients who are likely to show faster vs. slower rates of decline over time. This information in turn has potential to provide additional therapeutic insight by estimating the efficacy of disease-modifying therapies to “flatten” the slope of decline in each individual patient.

While the prognostic utility of tau PET signal in symptomatic stages of AD has been recently acknowledged^69^, there is limited evidence in this regard based on individuals presenting with atypical features of the disease, including non-amnestic clinical presentations and/or age of onset younger than 65 years. Patients with atypical AD tend to exhibit longitudinal clinical decline at a faster rate than those with typical AD^70–73^, highlighting the need for developing a reliable prognostication tool for patients with atypical early AD. In the present study, we investigated the role of tau PET signal localized to the default mode network at baseline in predicting clinical decline approximately one year later in patients at early clinical stages of atypical AD. We additionally compared the prognostic utility of baseline tau burden to that of cortical thickness and amyloid burden, all measured within the default mode network at baseline. Consistent with our hypotheses, we found that baseline tau in the default mode network was the strongest predictor of subsequent clinical decline, outperforming baseline cognitive and functional impairment, tau accumulation in other functional networks of the cerebral cortex, and atrophy and amyloid burden in the default mode network.

At baseline, early AD patients exhibited prominent tau accumulation in posterior cortical regions, including the major nodes of the default mode network (e.g., posterior cingulate, precuneus, angular gyrus, and lateral temporal cortex), with only modest involvement of the medial temporal and frontal regions, compared with Aβ-age-matched cognitively unimpaired control participants. These observations are overall consistent with prior work from our group^9,10,24,25,46^ and others^7,26,74^ identifying similar patterns of tau deposition in patients with variants of atypical AD. Aside from the default mode network, relatively higher tau signal was also identified in the dorsal attention and frontoparietal networks. This may reflect the high number of patients with a clinical diagnosis of PCA or amnestic dysexecutive AD in our study sample, where these functional networks are known to be particularly vulnerable^24,75,76^.

Next, we examined how baseline tau burden would predict subsequent clinical decline, as measured by the annualized change in CDR-SB scores from baseline to follow-up. Despite the popularity of the CDR scale as a primary endpoint in clinical research and trials for AD, to our knowledge there are only two published studies to date that have conducted tau-based prognostication of longitudinal clinical decline using this outcome measure. In one study, the authors examined a sample of patients with early AD and moderate AD as well as cognitively unimpaired participants and showed that greater baseline tau burden was associated with faster decline on CDR-SB, regardless of whether tau was defined at the level of whole cortical gray matter, temporal meta ROI, or Braak staging ROIs^21^. In another study of early AD, the authors failed to identify a significant relationship between baseline cortical tau and CDR-SB change^22^. While our patient sample and that of La Joie et al.^22^ are comparable in the level of baseline clinical impairment and the magnitude of decline over time, the two studies can be distinguished by the range of FTP PET signal (average cortical SUVR from ∼1 to ∼2.7 in La Joie et al. vs. 1.24 to 6.16 here), which may explain this discrepancy. The present study expands on prior findings by providing evidence that network-specific measures of tau accumulation can be used as a meaningful biomarker for disease progression in a clinically heterogeneous sample of patients with atypical early AD.

Our analysis demonstrated that the spatial topography of baseline tau deposition predicting subsequent clinical decline was much more widespread than that of group-average baseline tau deposition itself, which showed remarkable spatial overlap with the canonical default mode network. Current models of tau propagation in AD emphasize the posterior-to-anterior trajectory, such that as symptomatic AD progresses, tau spreads from posterior cortical areas to prefrontal cortex, including the nodes of the default mode network^26,38^. However, we found that the strength of the relationship between baseline tau and future clinical decline was comparable across the four lobar ROIs within the network. This suggests that tau accumulation in both the posterior nodes (where tau likely originates in the network) and anterior nodes (to which tau likely spreads in the network) of the default mode network are similarly important in determining the future rate of clinical decline in atypical early AD.

Further underscoring the unique contribution of the default mode network, we found that baseline tau burden in this network predicts subsequent clinical decline more strongly than that in any other functional network of the cerebral cortex. In fact, simply including baseline tau in the default mode network as a covariate in addition to age, sex, and CDR-SB scores at baseline more than tripled the amount of explained variance in the data. Moreover, our data-driven model selection approach revealed that a model with baseline tau in the default mode network as the sole predictor provides fit as good as the best performing model with the lowest AIC. These findings suggest that the prognostic utility of tau PET signal is not similar across networks and that the default mode network shows maximal sensitivity to predicting future clinical decline in early AD.

Our analysis of additional imaging biomarkers showed that baseline cortical thickness of the default mode network estimated from MRI also predicts subsequent clinical decline, although to a lesser extent than baseline tau burden in the same network; amyloid deposition in the default mode network did not predict subsequent clinical decline. When these imaging biomarkers were considered together in multimodal prediction models, one of the most parsimonious models included baseline tau and cortical thickness in the default mode network as the only predictors, suggesting complementary contributions of these measures to predicting clinical decline in atypical early AD. These findings are in line with previous investigations on biomarker-based prognostication in AD, which have provided converging evidence that tau PET is a superior predictor of cognitive decline compared with other measures of AD pathology and neurodegeneration including amyloid burden and gray matter atrophy in symptomatic AD patients^11–14,16–20^. This pattern of brain-behavior associations is also consistent with prior cross-sectional work by us and others. For example, in a recent study of PCA patients we showed that tau burden within the dorsal attention network was associated with visuospatial cognitive impairment while controlling for cortical thickness in the same network^24^. Partially independent associations of tau and cortical atrophy with cognition have been similarly reported across the syndromic spectrum of AD^77–79^. In a series of studies, we have also demonstrated the utility of cortical thickness as a biomarker of current clinical impairment or future clinical decline across AD phenotypes^80–83^. Although the magnitude of cortical atrophy was less prominent than that of tau burden in our patient sample, the present results suggest that modeling cortical thickness in addition to tau helps better predict subsequent clinical decline, likely by capturing aspects of early neurodegenerative processes following tau deposition.

Finally, as a first step toward developing a tool for individualized risk stratification, we compared baseline characteristics between a subset of our patients exhibiting faster clinical decline and the rest of the sample. Consistent with our findings discussed so far, we found that early AD patients who went on to show clinical decline at a relatively faster rate had—at baseline— prominently greater tau burden and cortical atrophy in the default mode network, somewhat greater cognitive impairment, and modestly greater amyloid burden in the default mode network. Although we were limited in our ability to go beyond this simple analysis due to the sample size, these findings set the stage for future prognostic work aimed at providing individualized estimates of the likelihood of relatively more rapid clinical decline. This information has profound implications for patients, families, and professionals in making better-informed decisions about treatment, caregiving, and life planning.

Our study has some limitations that should be acknowledged, along with possible avenues for future research. First, we analyzed only baseline imaging data and clinical decline estimated from two time points approximately one year apart. Future work should examine data acquired from more time points to reduce the possibility of measurement error. Longitudinal neuroimaging would also be useful in more comprehensively characterizing dynamic trajectories of AD-related neuropathologic changes, neurodegeneration, clinical decline, and the relationships between these phenomena. Second, while we purposefully focused on patients with early AD, we may be able to gain a better understanding of the prognostic utility of network-specific tau PET signal by studying samples with tighter ranges of clinical impairment at baseline (by focusing solely on MCI, for example). An overall larger sample would also be useful in minimizing the risk of model overfitting in future work. Third, we used the same ROI mask to define the default mode network in each patient based on an established parcellation of the cerebral cortex. While this is a common analytical approach in the literature, it is important to acknowledge potential variability in the topography of functional networks across individual patients^84,85^. A recent tau PET study in AD has also demonstrated that participant-specific ROIs show greater sensitivity to detecting longitudinal tau spread than group-level ROIs^86^. Future work should compare individual-specific and group-based approaches to neuroimaging data analyses to see how much this distinction influences clinical prognostication.

In sum, the present findings support the strong and specific contribution of baseline tau burden within the default mode network of the cerebral cortex to predicting the magnitude of clinical decline in a sample of atypical early AD patients one year later. This simple measure based on a tau PET scan could aid the development of a personalized prognostic, monitoring, and treatment plan tailored to each individual patient, which would help clinicians not only predict the natural evolution of the disease but also estimate the effect of disease-modifying therapies on slowing subsequent clinical decline given the patient’s tau burden while still early in the disease course.

## Data Availability

The data that support the findings of this study are available from the corresponding authors, upon reasonable request.

## Acknowledgements

The authors would like to thank the patients and families who participated in this research, without whose partnership this research would not have been possible.

