## Supplementary Methods for "Default mode network tau predicts future clinical decline in atypical early Alzheimer’s disease"

### Supplementary Methods: Calibration of amyloid PET SUVR data to Centiloid units

In the current patient sample, baseline amyloid deposition was quantified using FBB PET for 21 patients, whereas PiB PET was used for 27 patients. To harmonize the data across these different radioligands, we converted both FBB and PiB SUVR images to Centiloid (CL) units following the published guidelines[1,2]. Individual FBB PET frames were first spatially aligned and averaged. Following the ADNI protocol for PET data preprocessing, we resampled each participant's mean FBB PET image to 1.5 mm isotropic voxels. We then estimated the intrinsic smoothness of each participant's FBB PET image via 3dFWHMx in AFNI[3,4]. The estimated FWHM parameters were used to calculate the smoothing filter needed to yield the desired FWHM of 8 mm[5] and the resulting FBB PET image was co-registered to native space MRI data using SPM12 for each participant. We calculated a neocortical composite FBB SUVR based on FreeSurfer-derived segmentations of frontal, temporal, parietal, and cingulate regions with the whole cerebellum as a reference region. Finally, FBB SUVR data were converted to CL units using the published equation (equation 5 in Royse et al.[2]).

To calibrate PiB PET data to CL units, we first downloaded the *Standard PiB dataset* from the Global Alzheimer's Association Interactive Network (GAAIN) (https://www.gaain.org/centiloid-project), consisting of structural MRI and PiB PET data acquired 50 to 70 min post-injection from a sample of 45 AD patients and 34 cognitively unimpaired young adult participants[1]. We processed these data with a local implementation of the Standard Centiloid preprocessing (STD) pipeline based on SPM8 as described in Klunk et al.[1], which produced mean SUVR values in a large composite cortical region using whole cerebellum as a reference region ($\text{PiB\_STD\_WC}_{\text{SUVR}}$). We then compared these locally replicated SUVR values with the original SUVR data published by Klunk et al.[1] using the same dataset and pipeline. To pass the quality control, replicated SUVR values must fall within 2% of the group means for both AD patients (2.076 ± 2% = [2.056, 2.096]) and young control participants (1.009 ± 2% = [0.99, 1.029]) originally reported by Klunk et al.[1]. Our results fully met these criteria (AD group mean: 2.086 ± 0.204, control group mean: 1.014 ± 0.047), demonstrating a successful implementation of the STD pipeline. We then used these replicated group means to convert SUVR data to CL units ($\text{PiB\_STD\_WC}_{\text{CL}}$) with the following equation.

$$\text{PiB\_STD\_WC}_{\text{CL}} = 100 \times \frac{(\text{PiB\_STD\_WC}_{\text{SUVR}} - 1.014)}{(2.086 - 1.014)} \qquad (1)$$

Replicated CL values ($y$) were then regressed against the original CL values reported by Klunk et al.[1] ($x$) to obtain the slope, intercept, and $R^2$. The observed regression parameters all met the criteria for quality control: Slope = 1.002 (must be between [0.98, 1.02]), intercept = -0.107 (must be between [-2, 2]), and $R^2$ = .999 (must be > .98) (**Supplementary Fig. 1A**).

Next, we processed the *Standard PiB dataset* via a FreeSurfer-based pipeline described above. We then performed linear regression of the resulting SUVR values ($\text{PiB\_FS\_WC}_{\text{SUVR}}$) ($y$) on the SUVR values obtained the STD pipeline ($x$) to confirm that our pipeline produces reliable results relative to the standard method. The relationship between these two sets of PiB SUVR data was very strong ($R^2$ = .987), fulfilling the Centiloid criteria for reliability ($R^2$ > .70)[1], supporting the reliability of our data processing pipeline (**Supplementary Fig. 1B**). Using the slope and intercept obtained here, we transformed $\text{PiB\_FS\_WC}_{\text{SUVR}}$ values calculated from a subset of our patients ($n$ = 27) to estimated SUVR values from the STD pipeline ($^{\text{calc}}\text{PiB\_STD\_WC}_{\text{SUVR}}$), with the following equation:

$$^{calc}PiB\_STD\_WC_{SUVR} = \frac{\text{PiB\_FS\_WC}_{\text{SUVR}} - 0.214}{0.749} \qquad (2)$$

These estimated standard SUVR values were then converted to CL units using Eq. 1 above. Finally, for each patient we generated a vertex-wise CL parametric image by applying the conversion factors derived from the analysis described above to their respective vertex-wise maps of $FBB\_FS\_WC_{SUVR}$ and $^{calc}PiB\_STD\_WC_{SUVR}$.

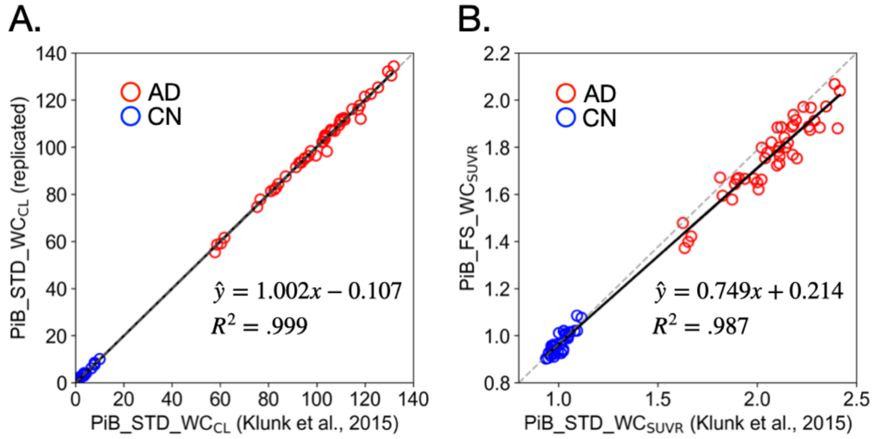


**Supplementary Figure 1. Calibration of amyloid PET data for differences in radiotracer and processing pipeline.** (A) Successful replication of PiB PET data (expressed as Centiloid [CL] units) in a sample of AD patients (*n* = 45) and cognitively unimpaired control participants (*n* = 34) via a local implementation of the standard PiB processing pipeline as described by Klunk et al.[1]. (B) Validation of a custom data processing pipeline based on FreeSurfer regions of interest (see Materials and Methods) using the same sample.

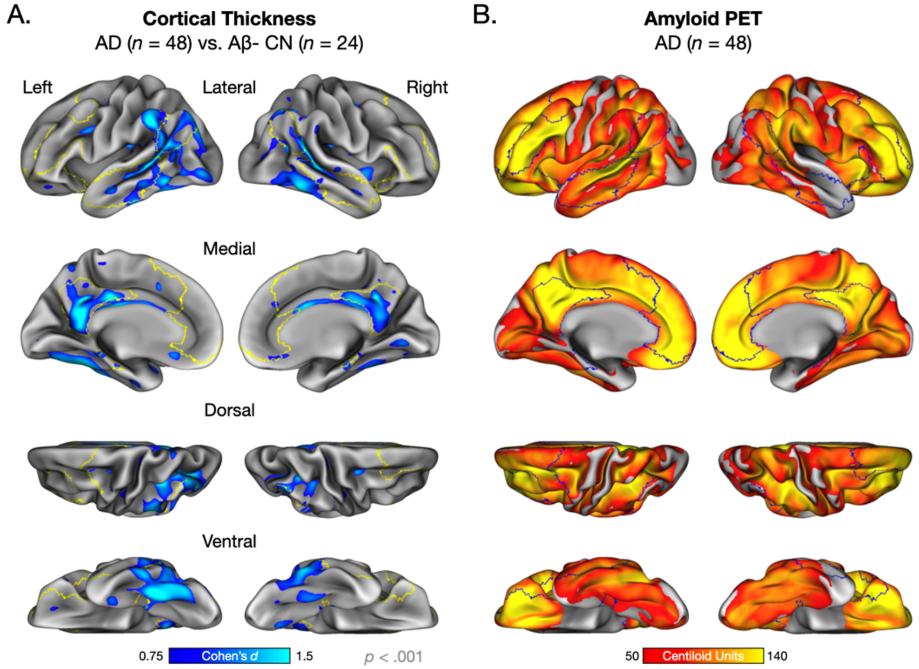


**Supplementary Figure 2. Abnormal cortical thickness and amyloid PET signal in early atypical Alzheimer's disease.** (A) Colored vertices on the cortical surface map indicate areas where early Alzheimer's disease (AD) patients ($n$ = 48) had reduced cortical thickness than Aβ- cognitively normal participants ($n$ = 24). Statistical significance was assessed at vertex-wise $p < .001$. The DMN is outlined in yellow. (B) The cortical surface map identifies the magnitude of amyloid PET tracer uptake expressed as Centiloid units. This map was created by a series of linear regression to calibrate for differences in radiotracer (PiB or FBB) following the published guidelines (see Materials and Methods).

**Supplementary Table 1. Baseline characteristics of early AD patients exhibiting clinical decline at faster than average vs. average rate.**

| | Faster than average (>1 *SD*) | Average (≤1 *SD*) |
|---|---|---|
| *n* | 7 | 41 |
| Age (years) | 63.59 ± 6.51 | 64.2 ± 8.47 |
| Sex (M/F) | 1/6 | 19/22 |
| Education (years) | 16.88 ± 2.57 | 15.71 ± 1.38 |
| CDR-SB | 5 ± 2.8 | 3.27 ± 1.57 |
| FTP DMN (SUVR) | 4.3 ± 1.37 | 3.07 ± 1.13 |
| Cortlcal Thickness DMN (mm) | 2.24 ± 0.12 | 2.36 ± 0.12 |
| Amyloid DMN (CL) | 114.56 ± 34.21 | 105.71 ± 25.07 |

CDR-SB = Clinical Dementia Rating Sum-of-Boxes; FTP = $^{18}$F-Flortaucipir PET; DMN = default mode network; CL = Centiloid units.

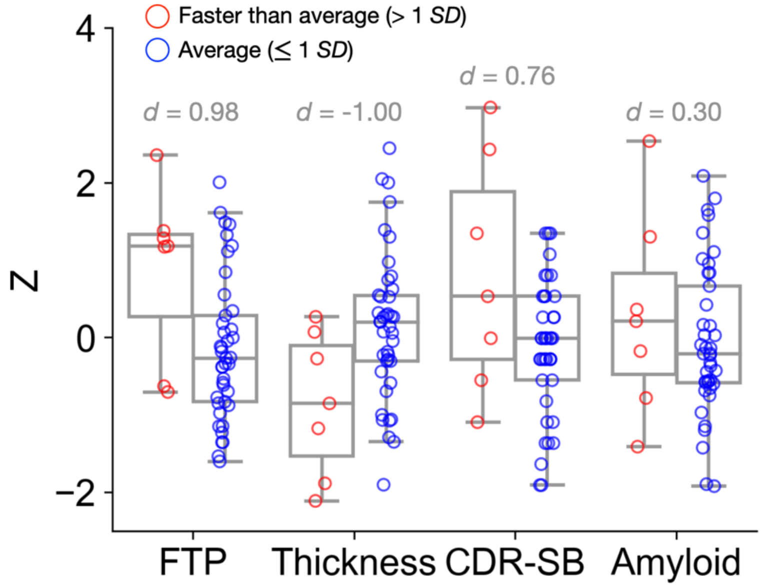


**Supplementary Figure 2. Baseline imaging and clinical characteristics of early AD patients exhibiting clinical decline at faster than average vs. average rate.** Box plots represent the median, range, and distribution of baseline $^{18}$F-Flortaucipir (FTP) PET signal, cortical thickness, and amyloid PET signal within the default mode network as well as CDR-SB scores, separately for those patients showing longitudinal clinical decline at faster than average (*n* = 7) and average rate (*n* = 41). All data are shown as *Z*-scores. Values shown in gray indicate Cohen's *d* effect size for each group comparison. CDR-SB = Clinical Dementia Rating Sum-of-Boxes.
